# Modelling the effectiveness of isolation strategies for managing mpox outbreaks with variable infectiousness profiles

**DOI:** 10.1101/2023.10.04.23296551

**Authors:** Yong Dam Jeong, Takara Nishiyama, Hyeongki Park, Masahiro Ishikane, Noriko Iwamoto, Kazuyuki Aihara, Koichi Watashi, Eline Op de Coul, William S Hart, Robin N Thompson, Norio Ohmagari, Jacco Wallinga, Shingo Iwami, Fuminari Miura

## Abstract

The global outbreak of mpox (formerly monkeypox) in 2022 raised public awareness about the disease. The ensuing sporadic outbreaks in 2023 highlighted the importance of sustaining nonpharmaceutical interventions, such as case isolation and contact tracing. Using viral load data, we developed a modelling framework to characterize the various infectiousness profiles of infected individuals. We used this model to examine the potential effectiveness of two different possible isolation rules: specifically, rules permitting infected individuals to stop isolating after either a fixed-duration or following negative tests for infection. Our analysis showed large individual variations in the duration of viral shedding, ranging from about 23 to 50 days. The risk of infected individuals ending isolation too early (i.e., while they remained an infection risk) was estimated to be about 5% after 3 weeks of isolation. Unnecessary isolation after the end of the infectious period could be reduced by use of a testing-based rule. These findings support the choice of a 3-week isolation period following symptom onset if a fixed-duration rule is used, but also demonstrate how testing can mitigate unnecessarily prolonged isolation for those who have shorter infectious periods.

## Main Text

Since May 2022, a global outbreak of mpox (formerly monkeypox) has spread primarily among men who have sex with men (MSM), first in European and North American countries, and later in other regions^1^. Although growth of the outbreak was initially rapid, the global trend in reported cases changed around the summer of 2022 and has been declining ever since^2^. Recent studies have suggested that the case saturation in many countries may be explained largely by infection-derived immunity accumulated among individuals who have many sexual partners^3^, and the following decline in cases may have been accelerated by vaccination campaigns or behavioral changes^4,5^. However, since the beginning of 2023, sporadic outbreaks have been reported, mainly in Asian countries, which were least affected by the 2022 outbreak and where vaccination campaigns had not yet been initiated, leading to a substantial number of individuals remaining at risk of infection^6^. There are also reports of some breakthrough infections and reinfections in European countries^7,8^. These findings warrant caution against a resurgence of mpox and highlight the importance of maintaining nonpharmaceutical interventions (NPIs).

One essential NPI is case isolation. The effectiveness of isolation has been extensively studied in the context of COVID-19^9,10^ and other diseases^11,12^. In general, if cases are detected earlier (e.g., via contact tracing) and isolation begins sooner, a larger proportion of onward transmissions can be prevented^13^. When determining the end of isolation, later is always safer, as a longer isolation period minimizes the risk of releasing individuals who remain infectious. However, case identification may require a substantial public health investigation effort, and redundant isolation induces societal cost and burden for individuals^14,15^. Furthermore, more stringent control strategies may also lead to reduced efficacy due to non-adherence^16^. To mitigate these burdens, testing-based rules were applied during the COVID-19 pandemic^17,18^, which typically involved ending isolation following a specified number of successive negative PCR or antigen test results. Individual-level viral load data have been used to balance the effectiveness and cost of isolation rules^19^, which is key to sustainable implementation.

Current guidelines for mpox generally suggest individuals who are exposed to the virus quarantine for about three weeks^18,20-23^ and refrain from sexual contact for 12 weeks after the end of isolation^20^. The three-week monitoring period is based on the estimated incubation period^24,25^, in which more than 98% of cases show symptoms within 21 days of exposure. Although the exact duration of the infectious period is unclear, several studies have investigated serial intervals (i.e., the time interval between symptom onset dates of primary and secondary cases) and have suggested that, for symptomatic cases, more than 90% of transmission occurs within two weeks of symptom onset^26,27^. Recent findings have also indicated that there might be considerable heterogeneity in the infectious period between infected individuals. For example, there is substantial variation in observed serial intervals^26^, and some confirmed cases in Europe exhibited prolonged viral shedding in their bodily fluids^28,29^. Consequently, isolation rules that do not account for heterogeneity between infected individuals (e.g. rules based on isolating for a fixed period following symptom onset) may lead to either a risk of ending isolation too early for those who are still infectious, or an unnecessarily long isolation period for those who are no longer infectious.

In this study, we first characterize individual infectiousness profiles among mpox cases by analyzing longitudinal viral load data. We describe the time course of virus shedding using a mathematical model that captures individual heterogeneity in the duration of viral clearance. We then stratify the population according to the characterized shedding profiles and evaluate the effectiveness of two different isolation rule types: a fixed-duration rule and a testing-based rule (i.e., a tailored rule for individuals who test negative several times). Our study provides an approach to quantify both the risk of ending isolation too early and the period for which infected individuals are isolated but do not pose an infection risk. Our model can be used by policy makers to inform decision making, allowing isolation strategies to be determined that balance cost and effectiveness appropriately.

## Results

### Analyzed data and model fitting

We conducted a literature review of individual-level mpox patient data and identified a total of 90 mpox cases with lesion samples meeting the inclusion criteria (see **Methods**). All cases were symptomatic, and most of them were reported in Europe. To standardize the collected data, we converted the reported cycle threshold values to viral load (copies/ml) using the conversion formula proposed in a previous study^30^ (**Supplementary Table 1**). We then fitted the viral clearance model to the longitudinal viral load data from lesion samples (**Extended Data Fig. 1a** and **Supplementary** Fig. 1). Estimated parameters suggested a median viral load of 10^7.7^ copies/ml (95% CI: 10^7.3^-10^8.2^) at symptom onset. Additionally, a prolonged duration of viral shedding was estimated: the viral load dropped below the detection limit (10^2.9^ copies/ml) 30.9 days (95% CI: 23.4-50.6) after symptom onset (**Extended Data Fig. 1bc**). This finding is consistent with previous studies suggesting the persistent presence of mpox viruses in clinical specimens^28,31^.

### Stratification for mpox cases

The 90 analyzed mpox cases were stratified into three groups based on their estimated individual parameters using the K-means clustering algorithm: Group 1 (medium risk of transmission), Group 2 (high risk of transmission), and Group 3 (low risk of transmission) (**Fig. 1a** and **1b**): Group 3 was characterized by a lower viral load at symptom onset and faster clearance, whereas Group 2 was characterized by a higher viral load at symptom onset and slower clearance. In addition, to compare the viral dynamics between the three groups, we conducted statistical tests: Individuals in Group 2 had significantly higher viral loads at symptom onset than individuals in the other groups (*p* < 1.0 × 10^−3^ from the Mann-Whitney test). Also, individuals in this group had a larger area under the viral load curve (AUC) (*p* < 1.0 × 10^−3^ from the Mann-Whitney test), that is, the total amount of virus excreted between symptom onset and the end of shedding. Viral clearance was significantly faster in Group 3 than in the other groups (*p* < 1.0 × 10^−4^ from the Mann-Whitney test) (**Fig. 1c**). To describe the difference in timing of viral clearance, we additionally simulated the probability of detectable virus over time by using the model with estimated parameters for each group (**Fig. 1d**). In all stratified groups, the probability was greater than 90% at 3 weeks after symptom onset. However, in the total group (i.e., a group of all analyzed cases), the probability dropped to 69.9% (95% CI: 67.0-73.2) at 4 weeks after symptom onset, which is the upper bound of the isolation period recommended by the CDC^21^. The probability in Group 3 at 4 weeks after symptom onset was 55.3% (95% CI: 51.8-55.0), whereas the corresponding probability in Group 2 was 90.6% (95% CI: 88.7-92.1).

**Fig. 1:**
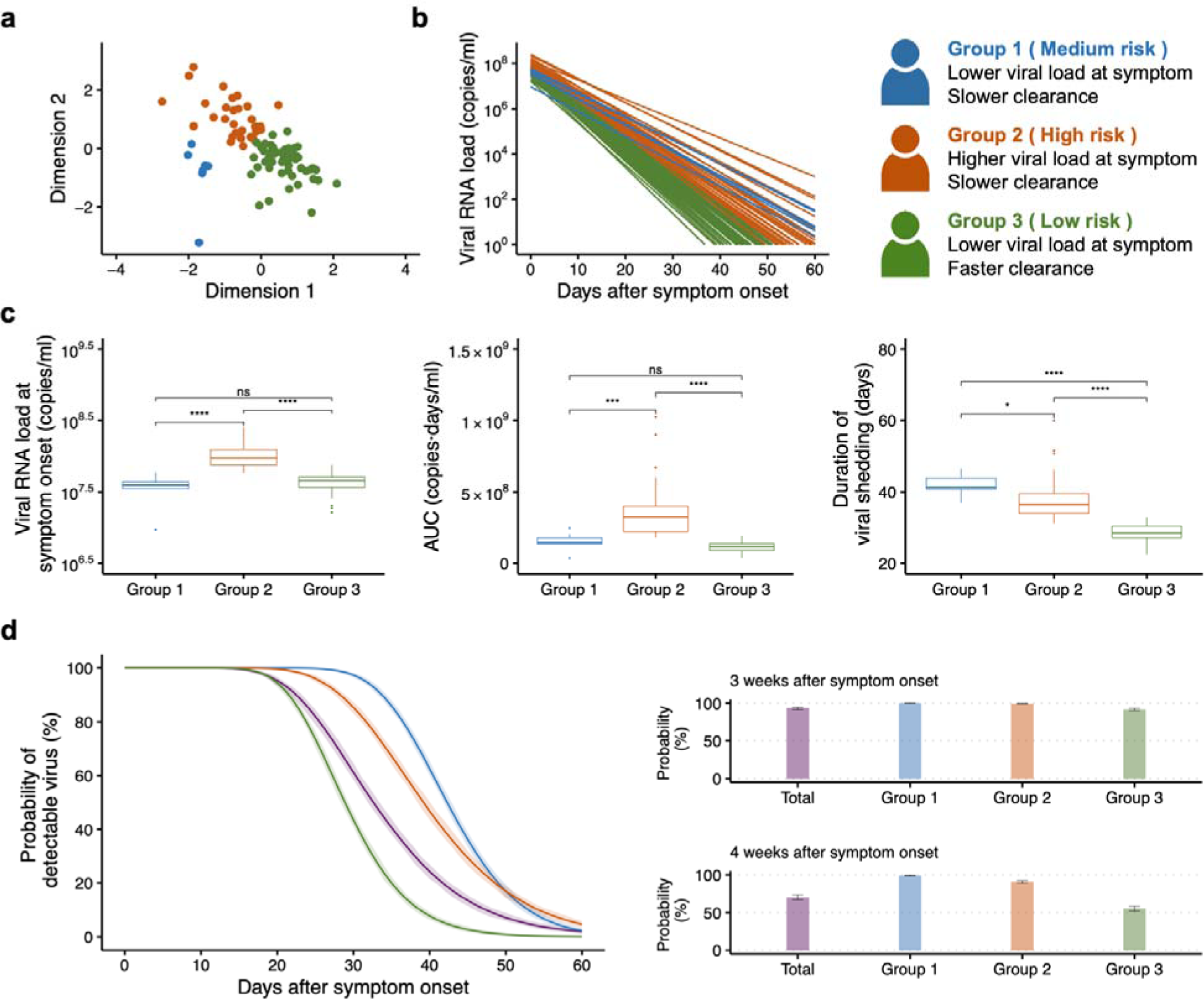
Stratification of mpox virus infections. **a,** Results of K-means clustering of mpox cases based on estimated individual parameters. Data points indicate individuals and are colored based on the group that each individual is in. The dimensions represent standardized estimated model parameters. **b,** Reconstructed individual mpox viral load trajectories in each group are shown. Group 1, Group 2, and Group 3 represent cases with medium, high, and low risk of transmission, respectively. **c,** Comparison between groups of: virus at symptom onset (left panel); area under the viral load curve (AUC), i.e., the total amount of virus shed over time (middle panel); and duration of viral clearance (right panel), respectively. The Mann-Whitney test was used to test for significant differences between groups. **d,** Viral clearance in each group. Probability of detectable virus after symptom onset for each group (left panel). The solid lines and shaded regions indicate means and 95% confidence intervals, respectively. Bar plots represent the probabilities for 3 weeks (right upper panel) and 4 weeks (right lower panel) after symptom onset, respectively.

### Fixed-duration rule

Under the estimated viral dynamics, we compared two types of rule for ending the isolation of mpox cases (fixed-duration and testing-based). To assess the effectiveness of the two rules, we considered three metrics: 1) the risk of prematurely ending isolation, 2) the average infectious period after ending isolation (where this period was defined to be zero for individuals who isolate beyond the duration of their infectious period), and 3) the average duration for which individuals were isolated unnecessarily after the end of their infectious period (which could be positive or negative). Whether an individual was infectious or not was ascertained based on a viral load threshold for infectiousness that was obtained from data on viral replication in cell culture^28,32,33^.

Under a fixed-duration rule of ending isolation 3 weeks after symptom onset, the risk of ending isolation prematurely in the total group was estimated to be 5.4% (95% CI: 4.1-6.7). The average duration for which individuals were isolated unnecessarily after the end of their infectious period was 8.3 days (95% CI: 8.0-8.6). Group 3 had a lower risk of ending isolation prematurely of 0.7% (95% CI: 0.3-1.3), and a longer unnecessary isolation period of 10.4 days (95% CI: 10.2-10.6). However, in Group 2, a higher risk of 16.1% (95% CI: 13.9-18.1) was estimated, with a shorter unnecessary isolation period of 4.6 days (95% CI: 4.2-4.9). To guarantee a risk of prematurely ending isolation below 5% and an infectious period after ending isolation shorter than 1 day, we found that the total group, Group 1, Group 2, and Group 3 needed to be isolated for 22, 23, 26, and 17 days, respectively. In this case, the duration for which individuals were isolated unnecessarily after the end of their infectious period was estimated to be 9.4, 8.1, 9.6, and 6.3 days for the total group, Group 1, Group 2, and Group 3, respectively (**Fig. 2a**).

**Fig 2:**
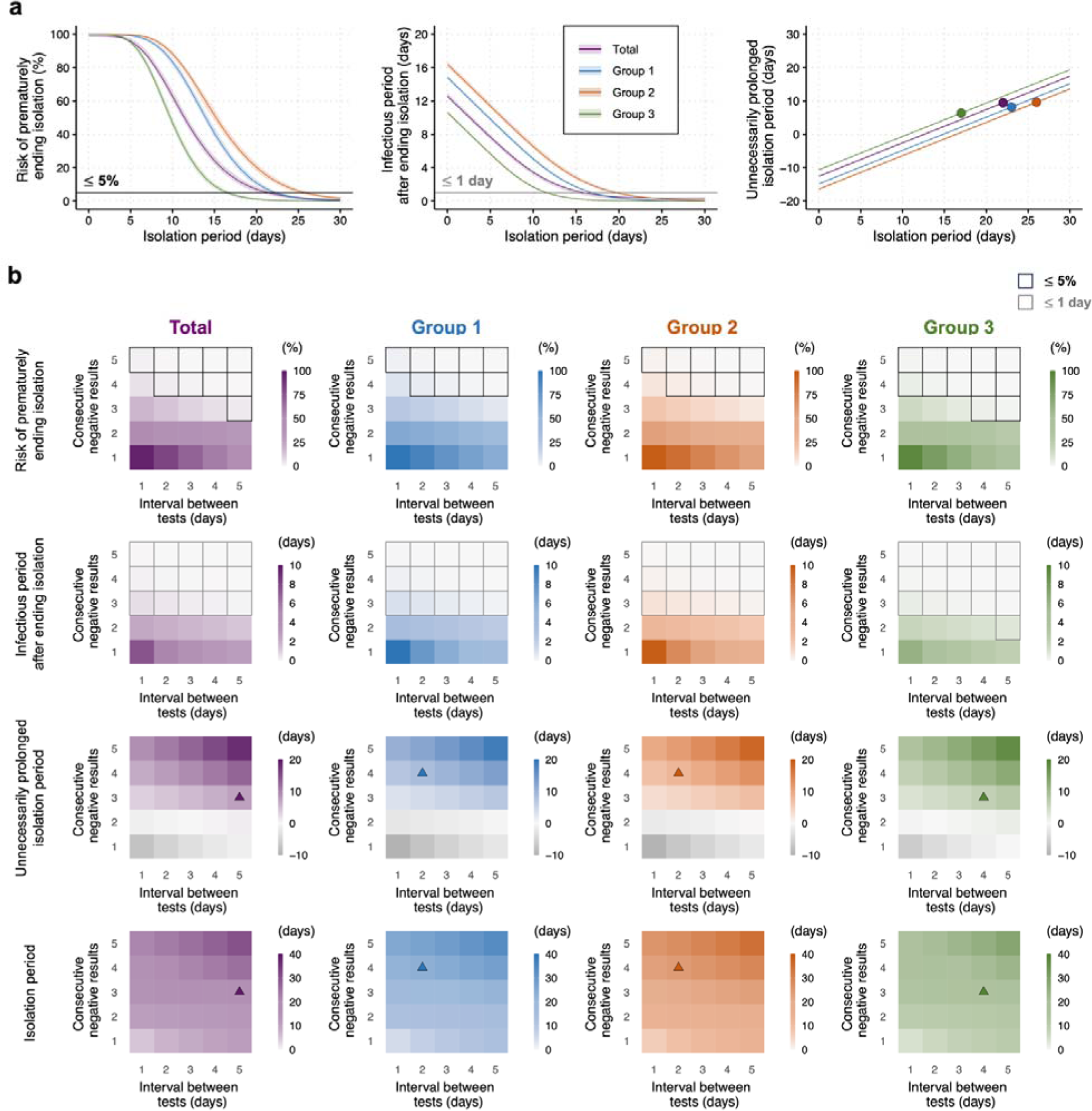
Fixed-duration and testing-based rules for different groups. **a,** Fixed-duration rules. The risk of prematurely ending isolation for different isolation periods (left panel). The black horizontal line corresponds to 5%. Infectious period after ending isolation for different isolation periods (middle panel). The grey horizontal line corresponds to 1 day. The time period for which individuals were isolated unnecessarily after the end of their infectious period for different isolation periods (right panel). The circles correspond to the points with the lowest unnecessarily prolonged isolation period for which the following conditions are satisfied: i) the risk of prematurely ending isolation was required to be lower than 5% and ii) the infectious period after ending isolation was required to be shorter than 1 day. The shaded regions in each panel indicate 95% confidence intervals. b, Testing-based rules. The risk of prematurely ending isolation (first row of panels), the infectious period after ending isolation (second row of panels), the isolation period following the end of infectiousness (third row of panels), and the optimal isolation period (fourth row of panels) are shown for different intervals between PCR tests and numbers of consecutive negative results necessary to end isolation. The areas surrounded by black and grey lines are those with 5% or lower risk of prematurely ending isolation and with 1 day or shorter infectious period after ending isolation, respectively. The triangles correspond to the points with the shortest isolation period following the end of infectiousness for which both conditions noted above are satisfied. Color keys and symbols apply to all panels. Note that the estimated values are based on 100 independent simulations.

### Testing-based rule

For a fixed-duration rule, isolation of mpox cases ends after a fixed time period following symptom onset, so the three metrics considered here can be calculated based on the isolation period alone (**Fig. 2a**). By contrast, a testing-based rule is dependent on both the time interval between tests and the exact criterion used for ending isolation (see **Methods**). Under a criterion in which isolation ends following two consecutive negative results with daily testing (a criterion widely used for COVID-19)^17^, the total group had a risk of prematurely ending isolation of 52.2% (95% CI: 49.7-54.6), and the infectious period after ending isolation was calculated to be 2.3 days (95% CI: 2.1-2.5). Similarly, high risks of prematurely ending isolation, accompanied with an infectious period after ending isolation longer than 1 day, were estimated in the stratified groups (first-row and second-row panels in **Fig 2b**).

By varying the criteria (i.e., the required number of consecutive negative results and the time interval between tests), different testing-based isolation strategies can be tested in terms of their effects on the three metrics. The risk of prematurely ending isolation and the infectious period after ending isolation decreased with a longer interval between tests and with a larger number of consecutive negative results (first-row and second-row panels in **Fig 2b**), whereas the duration for which individuals were isolated unnecessarily after the end of their infectious period increased (third-row panels in **Fig 2b**). Under the conditions that the risk of prematurely ending isolation is lower than 5% and infectious period after ending isolation is shorter than 1 day, the minimum value of the unnecessary isolation period in the total group was 7.4 days (95% CI: 7.1-7.7) with three consecutive negative results and an interval of 5 days between tests. Correspondingly, an isolation period of 20.1 days (95% CI: 19.7-20.5) was required on average (purple triangles in **Fig 2b**). On the other hand, under the same conditions, stricter strategies were needed for Group 2: four consecutive negative results and an interval of 2 days between tests were needed to minimize the duration for which individuals were isolated unnecessarily after the end of their infectious to 8.0 days (95% CI: 7.7-8.2), with a mean isolation period of 20.6 days (95% CI: 20.1-21.0) (red triangles in **Fig 2b**).

### Comparison between fixed-duration and testing-based rules for ending isolation

To highlight the difference between fixed-duration and testing-based rules for each group, we compared the two types of rule by computing the optimal isolation strategies in which the isolation period following the end of infectiousness is minimized while ensuring that the risk of prematurely ending isolation is less than 5% and the infectious period after ending isolation is less than 1 day. For testing-based rules, the mean isolation period that the optimal strategies then lead to can be computed for each stratified group. In the total group, the optimized fixed-duration and testing-based rules gave isolation periods of 22 and 20.1 days, resulting in minimized unnecessary isolation periods of 9.4 and 7.4 days, respectively (**Extended Data Fig. 2**). In Group 2, the testing-based rule led to an unnecessary isolation period that was 1.4 days shorter than the fixed-duration rule, whereas Groups 1 and 3 showed similar unnecessary isolation periods when either rule was applied. However, compared with the fixed-duration rule in the total group, the testing-based rule in Group 3 could reduce the optimal isolation period and the unnecessarily isolation period to 17.1 days and 6.5 days, respectively.

As a sensitivity analysis, we varied the assumed infectiousness threshold and investigated the corresponding difference in the period for which individuals were isolated unnecessarily after the end of their infectious period between the two rules (**Supplementary** Fig. 2). When the infectiousness threshold is higher, the corresponding infectious period becomes shorter, leading to a shorter required isolation period and shorter period of unnecessary isolation given the same acceptable risk. Our analysis showed that a higher infectiousness threshold resulted in smaller differences between the two rules for each stratified group (**Supplementary** Fig. 2), which was consistent with our previous findings for COVID-19^9^.

### Comparison between lesion and other samples for infectiousness after ending isolation

To demonstrate that lesion samples are suitable for designing isolation rules, we compared the viral dynamics that we inferred using lesion samples to analogous results obtained using other samples. Specifically, we used longitudinal viral load data measured in upper respiratory tract, blood, and semen samples from the same mpox cases to estimate mpox virus dynamics in those samples (**Supplementary Table 1** and **Extended Data Fig. 3a**). Following symptom onset, other samples exhibited lower viral loads compared with lesion samples. In particular, at the optimal ending isolation period of 22 days under a fixed-duration rule, the viral load in lesion samples was substantially higher than in other samples (**Extended Data Fig. 3b**). Moreover, we compared the predicted infectiousness when lesion samples and other samples are used by computing the proportion of individuals who remained infectious on day 22 after symptom onset. Around 3% of individuals were estimated to be infectious when lesion samples were used, whereas the viral load never exceeded the infectiousness threshold for the other samples (**Extended Data Fig. 3c**). This suggests that infectious mpox cases may be missed if we implement a testing-based rule with samples other than lesion samples.

## Discussion

In this study, we have compared different strategies that can be implemented to determine when mpox infected individuals stop isolating. If a single population-wide fixed-duration isolation strategy is used, then we found that allowing individuals to end their isolation after a period of three weeks following symptom onset is a reasonable threshold. Under this strategy, more than 95% of onward transmissions would be prevented. Our modelling analysis showed that there was individual heterogeneity in viral shedding kinetics, indicating that the use of testing-based rules may reduce the period for which infected individuals with a shorter duration of virus shedding are required to isolate.

We observed different shedding kinetics between patients in the analyzed data. We therefore stratified the patients based on their viral load during the decay phase of infection (**Fig. 1**). Variations in virus shedding may lead to substantial heterogeneity in infectiousness between individuals. For mpox, existing studies have focused on individual variations in the number of sexual contacts or partners^4,34^, because higher contact rates generally result in a larger number of secondary cases. In contrast, variations in viral shedding have received limited attention to date. Our study found that some mpox cases exhibit a 5–10-day shorter (or longer) duration of virus shedding than the average. Individuals in those groups may thus contribute to virus transmission for shorter (or longer) periods, resulting in a lower (or higher) number of secondary cases. When designing tailored interventions to ensure that the time-dependent reproduction number, *R* (the average number of secondary cases generated by each infected individual)^35^, is below one (i.e., the outbreak is declining), our approach provides a way to incorporate such heterogeneity in the infectious period by using individual viral load as a proxy.

For evaluating the risk of transmission following the end of isolation, the use of longitudinal viral load data may be advantageous over symptom-based approaches. One difficulty lies in the inherent uncertainty in self-reported symptoms; many confirmed mpox cases have been found to be not fully aware of their symptoms at the time of reporting^36^. Viral load data have the potential to provide more objective and quantitative criteria for ending isolation^37^. If viral load data are used, measuring viral load in lesion samples (rather than the other sample types that we considered) is the safest choice, as lesion samples showed the highest viral load and the longest detectable period (**Extended Data Fig. 3**).

Isolation rules need to balance the risk of releasing infectious cases prematurely and the societal burden of extended isolation. Our results indicate that optimized fixed-duration and testing-based rules can result in comparable risk levels if the same rule is applied to all cases. By contrast, the total duration of unnecessary isolation can be reduced using a testing-based rule. It is possible to shorten the isolation period for those with faster viral clearance, leading to a reduced burden at the population level. A testing-based approach may also be beneficial for evaluating the times at which individuals can resume sexual activities. Despite the current recommendation of using a condom for 12 weeks after scabs have fallen off as a precaution^20^, having additional information about patients’ infectiousness would offer reassurance to them and help to prevent discrimination and stigma related to sexual behaviors. As outlined in **Extended Data Fig. 3c**, our assessment revealed that viral load in semen had fallen below the infectiousness threshold by the endpoint of an optimized 22-day isolation period.

As with any modelling analysis, there are several limitations to our study. First, our analysis relied on the patient data collected, which may not be representative of all mpox cases in the affected population. The estimated parameters were obtained using data from untreated patients, but in outbreak settings antiviral drugs such as tecovirimat may be provided to infected individuals and used prophylactically. Further investigations into the association between clinical characteristics of patients, treatments, and viral load would be needed for a more granular understanding of heterogeneous infectiousness profiles. This may enable, for example, different fixed-duration isolation periods to be specified for individuals with different characteristics. Second, the association between mpox viral load and infectiousness needs to be understood more deeply. Specifically, based on experimental data on viral culturability, we used 10^6^ copies/ml as the infectiousness threshold value in main analysis; however, this involves uncertainty^28,32,33^. Accordingly, we performed sensitivity analyses (**Supplementary** Fig. 2) and found similar results regardless of the assumed threshold value. Last, our analysis did not capture external sources of uncertainty affecting the false-negative rate of testing. In practical settings, imperfect swab sampling, especially with self-collection, could occur (leading to differential test specificity/sensitivity). There may also be cases who repeatedly get tested until obtaining a negative result, as occurred during the COVID-19 pandemic^38^. Such challenges need to be considered when implementing testing-based rules.

In conclusion, this study provides empirical evidence for heterogeneity in virus shedding kinetics and infectiousness among mpox cases and describes the impact of such heterogeneity on the effectiveness of different isolation rules. Rules that recommend isolating following a fixed period after being exposed to the virus are straightforward to apply, and can be effective at preventing transmission. Rules that are instead based on obtaining negative test results prior to ending isolation are more nuanced and can have advantages, for example by reducing the period for which some individuals are required to isolate after they are no longer infectious. Careful consideration of the benefits and drawbacks of different strategies for ending isolation is essential. Ensuring sustainable implementation of NPIs remains key to responding effectively to future outbreaks of mpox.

## METHODS

### Viral load data

We searched the literature for longitudinal data from mpox cases meeting the following criteria: 1) viral load was measured at least at two different time points; 2) viral load was measured in different samples including lesion samples; and 3) patients did not receive any antivirals (as our model does not consider antiviral treatment). A total of 7 publications met those criteria, and 90 mpox cases were identified. We used only de-identified data from published studies and thus ethics approval was not required.

### Modelling mpox viral clearance and parameter estimation

Using the viral load data, we parameterized a mathematical model of temporal viral clearance dynamics in each infected individual. For this, we employed an exponential decay model, which was previously utilized in an mpox study^28^:

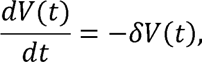

where the variable *V*(*t*) is the viral RNA load (copies/ml) at time *t* and parameter *δ* represents the viral clearance rate. Note that the timescale is time after symptom onset; *t* = 0 is thus the date on which symptoms of mpox first began. *V*(0) is the initial viral load at symptom onset.

A nonlinear mixed-effect model was used to estimate the parameters *δ* and *V*(0)^9,10^. This approach captures the heterogeneity in the viral dynamics by including both a fixed effect (the shared effect among individuals, i.e., population parameter) and a random effect (the individual-level effect) in each parameter. Population parameters and the standard deviation of random effects were estimated by using the Stochastic Approximation Expectation Maximization algorithm to compute the maximum likelihood estimator of the parameters, assuming a Gaussian distribution (mean 0 and variance σ^2^) for the residuals (i.e., differences between predicted log viral load and measured log viral load) to quantify the error used in our simulations^39^. Individual parameters were subsequently computed using Markov Chain Monte Carlo (MCMC). The estimation procedures were performed using MONOLIX 2023R1 (www.lixoft.com).

### Clustering algorithm to stratify mpox cases

We stratified the mpox cases using the K-means clustering algorithm^40^, which finds cluster assignments by minimizing the sum of squared Euclidean distances between estimated parameter sets. We first standardized the estimated parameters, since the two parameters in the viral clearance model have different units. Then, the algorithm partitioned the set of estimated individual parameters into *k* clusters. The optimal number of clusters was determined by the Silhouette method^41^.

### Simulation of viral dynamics and different rules for ending isolation

To account for individual variability in viral dynamics when determining the optimal duration of isolation, we simulated the predicted viral load, *V*(*t*), for 1000 virtual patients by running the viral clearance model. Parameter sets for each virtual patient were sampled from distributions of estimated model parameters. The measured viral load, *V*^^^(*t*), was obtained from the following equation: log_10_ *V*^^^(*t*) = log_10_ *V*(*t*) + *ε*, *ε* ∼ *N*(0, σ^2^), where *ε* is the error term. For each individual, the residuals were calculated at all measurement time points, and by fitting the Gaussian distribution to all computed residuals, the variance of error, σ^2^, was estimated. For parameter sets for virtual patients in each stratified group, we used individual parameters drawn from conditional distributions (i.e., distributions conditioned on the observed data and the estimated population parameters) characterizing the estimated parameters for individuals in each group, as estimated in the MCMC procedure.

Under a fixed-duration rule, it was assumed that isolation would end at a specified time following symptom onset. On the other hand, under testing-based rules using PCR tests, isolation ended when a given number of consecutive negative results was met with a given interval between tests. Here, we assumed that patients began to take tests immediately following symptom onset. To simulate various situations, we varied the interval between tests (from 1 to 5 days) and the number of consecutive negative results (from 1 to 5 times). To ascertain individuals’ infectious periods, a viral load threshold of infectiousness was considered. If the viral load of a patient was above the threshold, the patient was considered as being infectious. The threshold values considered were obtained from studies on viral replication in cell culture^28,32,33^. In this study, we set 10^6^ copies/ml as the main threshold value. However, this value is still uncertain and thus we also considered different threshold values from 10^5^ copies/ml to 10^7^ copies/ml as sensitivity analyses. For the evaluation of different rules, three metrics were computed: 1) risk of prematurely ending isolation, 2) infectious period after ending isolation, and 3) unnecessarily prolonged isolation period. The first metric gives the probability of releasing patients while they are still infectious. The second metric is defined as the mean number of days for which patients remain infectious after they are released from isolation (defined to be zero for an individual who is no longer infectious when released from isolation). The third metric gives the mean difference between the time when patients are no longer infectious and the time at which their isolation ends^9,10^ (which is negative for individuals who are infectious beyond the end of isolation). Specifically, the risk of prematurely ending isolation was computed as ∑_*i*_ *I*(*V*_*i*_(*τ*_*i*_) > Infectiousness threshold)/1000, where *I* is the indicator function, *V*_*i*_ is the predicted viral load of patient *i*, and *τ*_*i*_ is the time when isolation of patient *i* ends. We computed the infectious period after ending isolation by use of the following formula: ∑_*i*_ max (0, *τ*_*i*_ − *τ*_*i*_)/1000, where *τ*_*i*_ indicates the time when the predicted viral load of patient *i* drops below the infectiousness threshold. Finally, the unnecessarily prolonged isolation period was calculated as ∑_*i*_(*τ*_*i*_ − *τ*_*i*_)/1000. By running 100 simulations (1000 patients for each simulation), we reported the mean and 95% confidence intervals for distributions of those three metrics, respectively. All analyses were conducted using the statistical computing software R (version 4.2.3).

## LIST OF SUPPLEMENTARY MATERIALS

**Supplementary Fig. 1**: Estimated individual viral load trajectory for each mpox case with lesion samples

**Supplementary Fig. 2**: Comparison between fixed-duration and testing-based rules depending on different infectiousness thresholds

**Supplementary Fig. 3**: Estimated individual viral load trajectory for each mpox case with different samples

**Supplementary Table 1**: Summary of mpox viral load data with different samples

**Supplementary Table 2**: Estimated fixed effect parameters, standard deviation of random effect, and standard deviation of error in mpox viral loads for each sample

## Data Availability

All data produced in the present study are available upon reasonable request to the authors.

## ACKNOWLEDGMENTS

This study was supported in part by a National Research Foundation of Korea (NRF) grant funded by the Korea government (MSIT) (2022R1C1C2003637) (to K.S.K.); Scientific Research (KAKENHI) B 23H03497 (to S.I.); Grant-in-Aid for Transformative Research Areas 22H05215 (to S.I.); Grant-in-Aid for Challenging Research (Exploratory) 22K19829 (to S.I.); AMED CREST 19gm1310002 (to S.I.); AMED Research Program on Emerging and Re-emerging Infectious Diseases 22fk0108509 (to S.I.), 23fk0108684 (to S.I.), 23fk0108685 (to S.I.); AMED Research Program on HIV/AIDS 22fk0410052 (to S.I.); AMED Program for Basic and Clinical Research on Hepatitis 22fk0210094 (to S.I.); AMED Program on the Innovative Development and the Application of New Drugs for Hepatitis B 22fk0310504h0501 (to S.I.); AMED Strategic Research Program for Brain Sciences 22wm0425011s0302; AMED JP22dm0307009 (to K.A.); JST MIRAI JPMJMI22G1 (to S.I.); Moonshot R&D JPMJMS2021 (to K.A. and S.I.) and JPMJMS2025 (to S.I.); Institute of AI and Beyond at the University of Tokyo (to K.A.); Shin-Nihon of Advanced Medical Research (to S.I.); SECOM Science and Technology Foundation (to S.I.); The Japan Prize Foundation (to S.I.). The collaboration between R.N.T. and S.I. was supported by a Royal Society International Exchange award (grant number IES-R3-193037).

## AUTHOR CONTRIBUTIONS

SI and FM designed the research. YDJ carried out the computational analysis. SI and FM supervised the project. All authors discussed the research and contributed to writing the manuscript.

## EXTENDED DATA FIGURES

**Extended Data Fig. 1:**
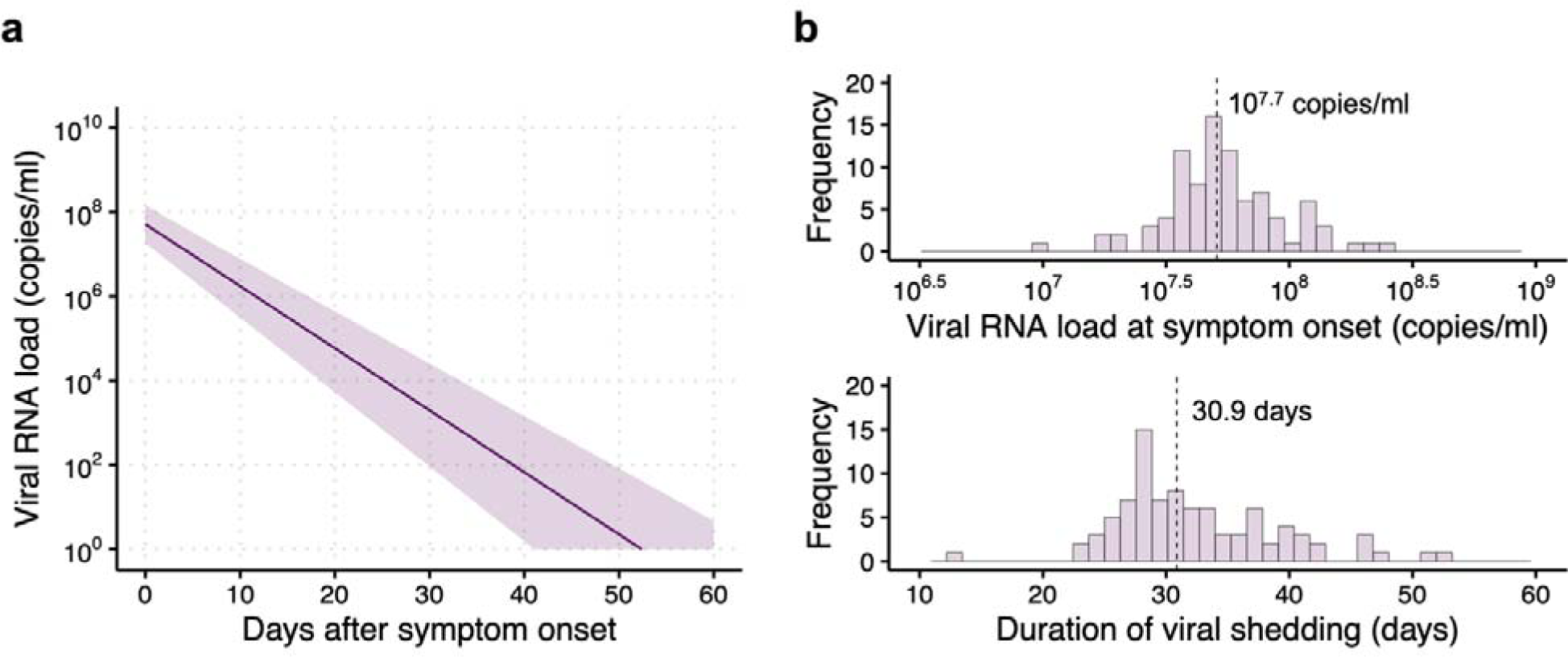
Characterization of mpox virus infection dynamics. **a,** Estimated viral load trajectory for mpox cases with lesion samples. The solid line is the estimated viral load trajectory under the best-fit fixed effect parameters. The shaded regions indicate 95% prediction intervals computed using a bootstrap approach. **b,** Distributions of estimates of the viral RNA load at symptom onset (upper panel) and distributions of estimates of the duration of viral shedding (lower panel). The vertical dashed lines indicate median values.

**Extended Data Fig. 2:**
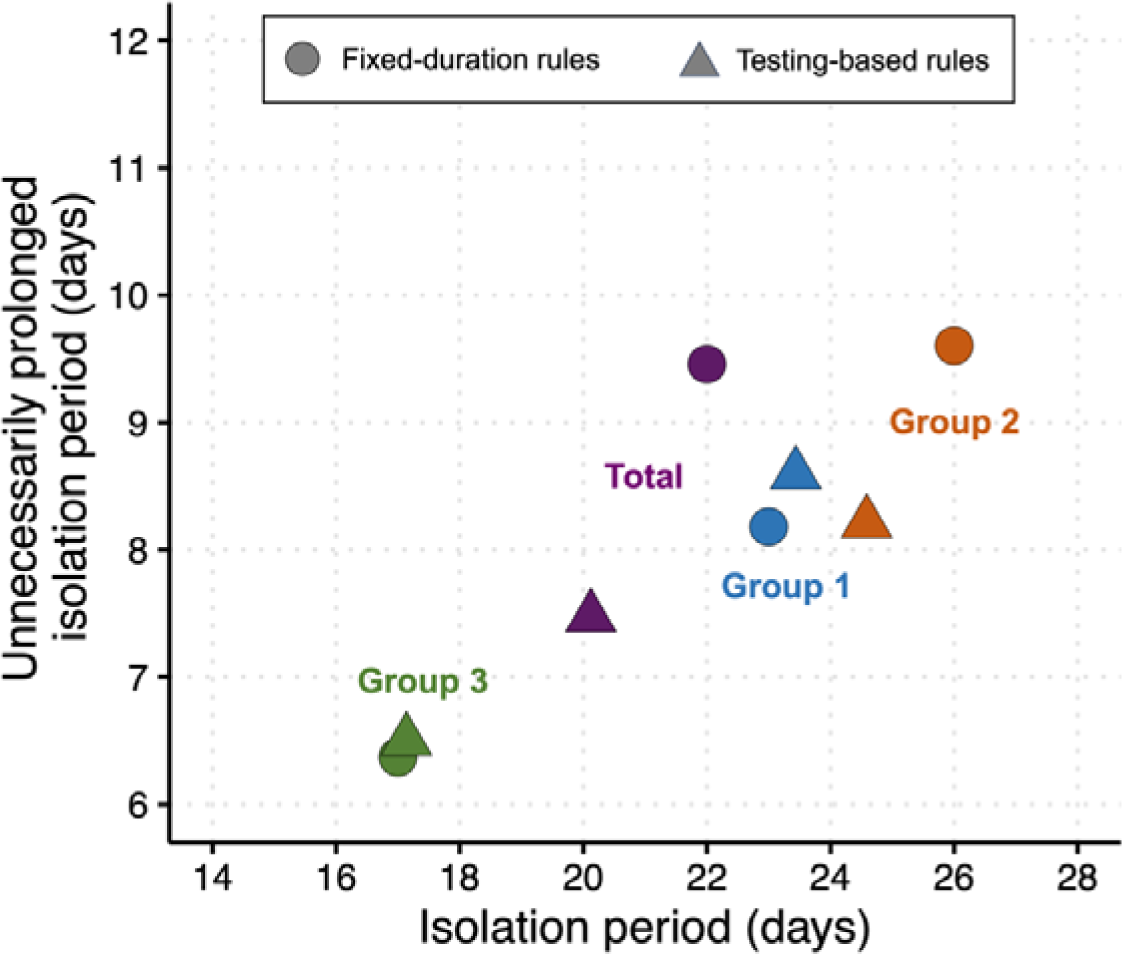
Comparison between fixed-duration and testing-based rules for different groups. The circles and triangles represent fixed-duration and testing-based rules, respectively. Each symbol represents the mean length of isolation using the strategy that minimizes unnecessarily prolonged isolation under the conditions that the risk of prematurely ending isolation is less than 5% and the infectious period after ending isolation is less than 1 day. Note that for testing-based rules, the interval between tests and the number of consecutive negative results necessary to end isolation were chosen to minimize the unnecessarily prolonged isolation period.

**Extended Data Fig. 3:**
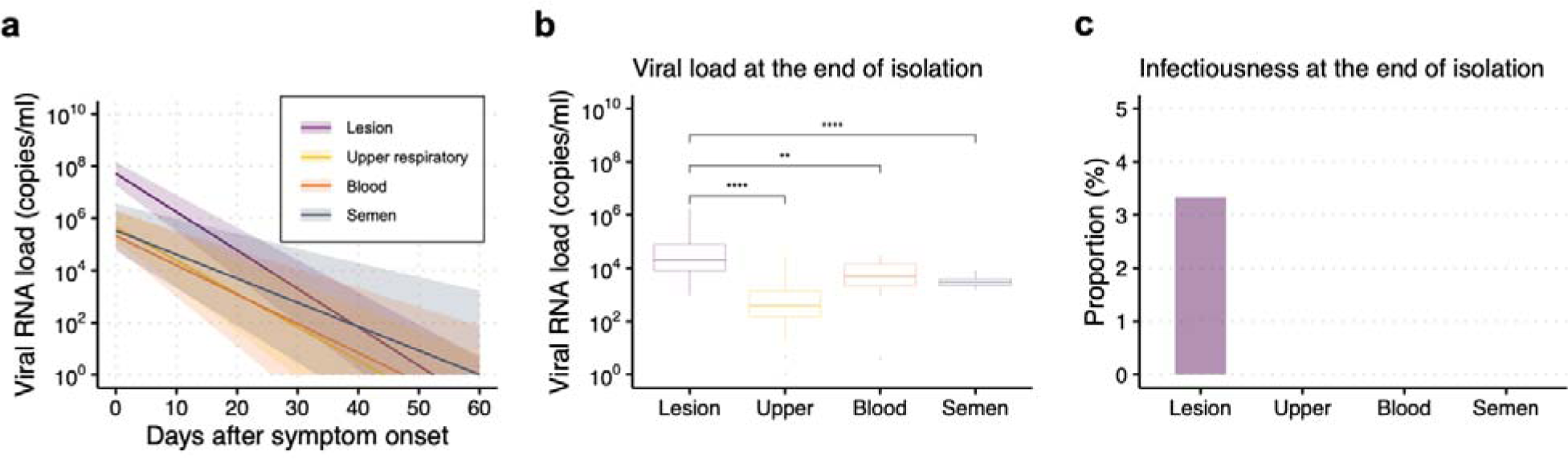
Comparison of mpox virus infection between lesion samples and other samples. **a,** Estimated viral load trajectories for mpox cases with different samples. The solid lines are estimated viral load trajectories for the best-fit parameters of fixed effect. The shaded regions indicate 95% prediction intervals computed using a bootstrap approach. The purple, yellow, orange, and grey colors correspond to lesion, upper respiratory, blood, and semen samples, respectively. **b,** Distribution of viral RNA load with different samples at the end of isolation. **c,** Proportion of infectious individuals with different samples (%) at the end of isolation. Note that the optimal ending isolation period of 22 days under a fixed-duration rule was used as the end of isolation. Color keys apply to all panels.

