## Supplementary Information for "Modelling the effectiveness of isolation strategies for managing mpox outbreaks with variable infectiousness profiles"

^1^interdisciplinary Biology Laboratory (iBLab), Division of Natural Science, Graduate School of Science, Nagoya University, Nagoya, Japan. ^2^Disease Control and Prevention Centre, National Centre for Global Health and Medicine, Tokyo, Japan. ^3^International Research Center for Neurointelligence, The University of Tokyo Institutes for Advanced Study, The University of Tokyo, Tokyo, Japan. ^4^Research Center for Drug and Vaccine Development, National Institute of Infectious Diseases, Tokyo, Japan. ^5^Centre for Infectious Disease Control, National Institute for Public Health and the Environment (RIVM), Bilthoven, the Netherlands. ^6^Mathematical Institute, University of Oxford, Oxford, OX2 6GG, UK. ^7^Department of Biomedical Data Sciences, Leiden University Medical Center (LUMC), Leiden, the Netherlands. ^8^Institute of Mathematics for Industry, Kyushu University, Fukuoka, Japan. ^9^Institute for the Advanced Study of Human Biology (ASHBi), Kyoto University, Kyoto, Japan. ^10^Interdisciplinary Theoretical and Mathematical Sciences Program (iTHEMS), RIKEN, Saitama, Japan. ^11^NEXT-Ganken Program, Japanese Foundation for Cancer Research (JFCR), Tokyo, Japan. ^12^Science Groove Inc., Fukuoka, Japan. ^13^Center for Marine Environmental Studies (CMES), Ehime University, Ehime, Japan.

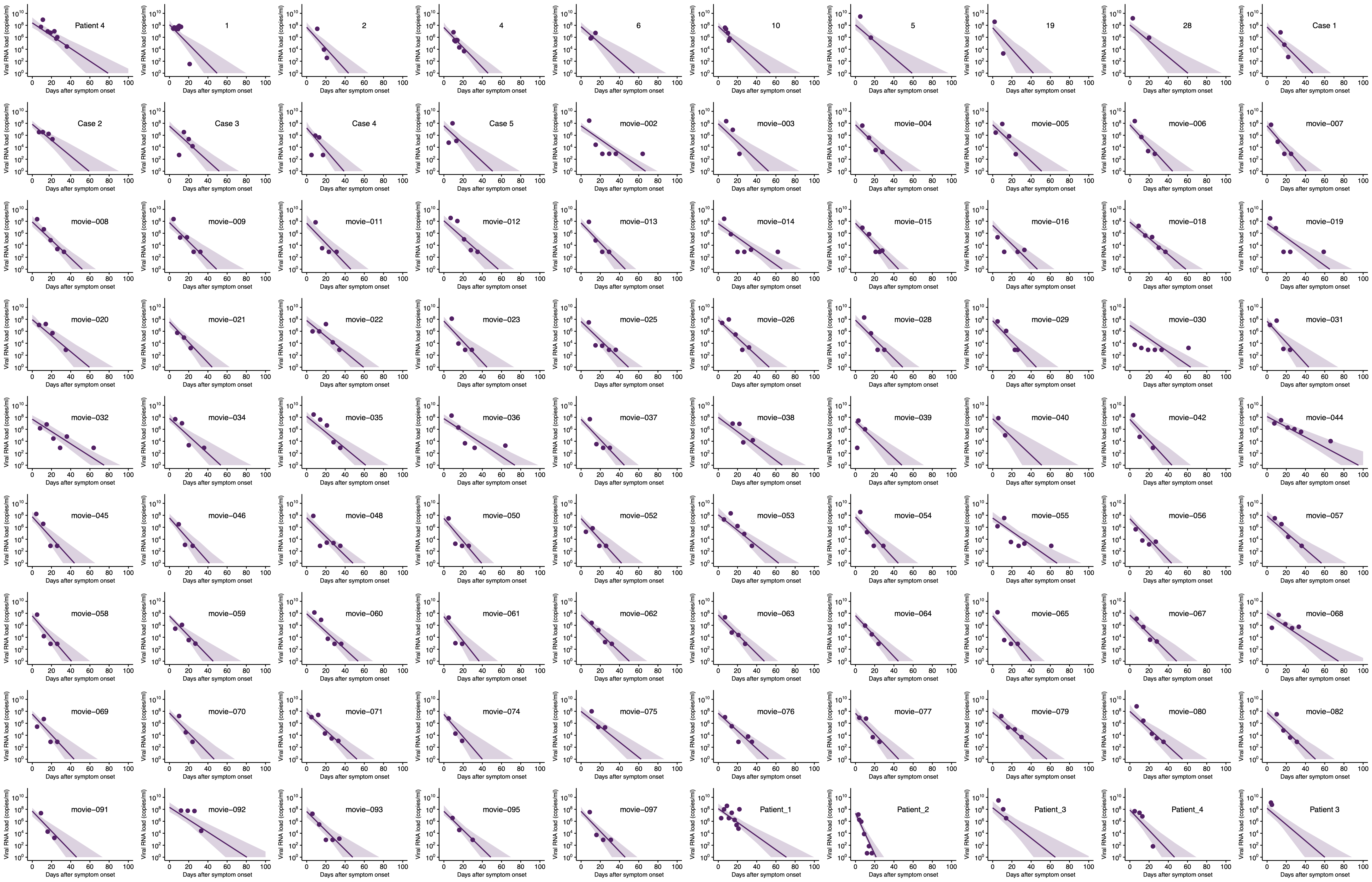

Supplementary Fig. 1: Estimated individual viral load trajectory for each mpox case with lesion samples. The solid lines are estimated viral load trajectories for the best-fit parameters. The shaded regions indicate 95% predictive intervals. The closed circles represent measured viral load observations.

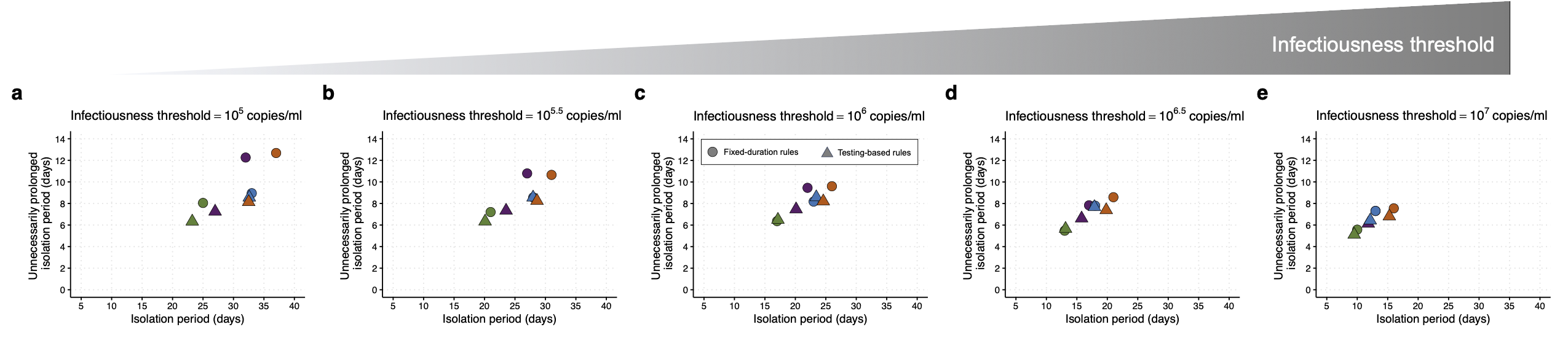

Supplementary Fig. 2: Comparison between fixed-duration and testing-based rules depending on different infectiousness thresholds. a, 10^5.0^ copies/ml. b, 10^5.5^ copies/ml. c, 10^6.0^ copies/ml. d, 10^6.5^ copies/ml. e, 10^7.0^ copies/ml. The circles and triangles represent fixed-duration and testing-based rules, respectively. Each symbol represents the mean length of the isolation period using the strategy that minimizes unnecessarily prolonged isolation under conditions that the risk of prematurely ending isolation is less than 5% and the infectious period after ending isolation is less than 1 day. Note that for testing-based rules, the interval between tests and the number of consecutive negative results necessary to end isolation were chosen to minimize the unnecessarily prolonged isolation period. The purple, blue, red, and green colors indicate total, Group 1, Group 2, and Group 3, respectively. Color keys and symbols apply to all panels.

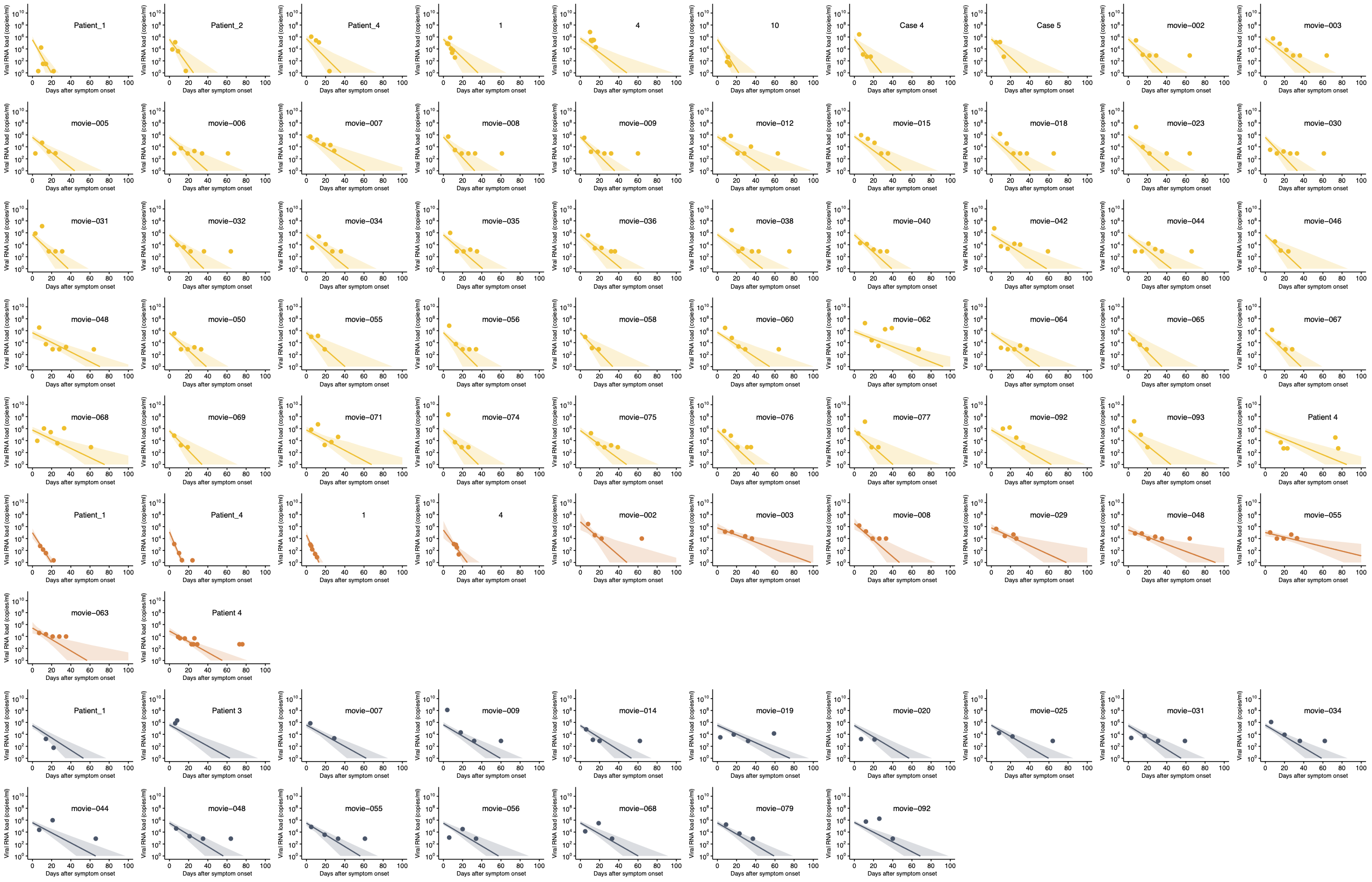

Supplementary Fig. 3: Estimated individual viral load trajectory for each mpox case with different samples. The solid lines are estimated viral load trajectories for the best-fit parameters. The shaded regions indicate 95% predictive intervals. The closed circles represent measured viral load observations. The yellow, orange, and grey colors correspond to upper respiratory, blood, and semen samples, respectively.

Supplementary Table 1: Summary of mpox viral load data with different samples

| **Samples** | **Number of data** | **Reporting unit** | **Source** |
| --- | --- | --- | --- |
| **Lesion** ( $n=90$ ) |  |  |  |
| Spain | $71$ | viral load (copies/ml) | ^1^ |
| Germany | $5$ | viral load (copies/ml) | ^2^ |
| UK | $1$ | cycle threshold^#^ | ^3^ |
| Italy | $3$ | cycle threshold^#^ | ^4^ |
| India | $5$ | cycle threshold^#^ | ^5^ |
| Germany | $4$ | cycle threshold^#^ | ^6^ |
| Italy | $1$ | quantification cycle^#^ | ^7^ |
| **Upper respiratory tract** ( $n=50$ ) |  |  |  |
| Spain | $41$ | viral load (copies/ml) | ^1^ |
| Germany | $3$ | viral load (copies/ml) | ^2^ |
| UK | $1$ | cycle threshold^#^ | ^3^ |
| India | $2$ | cycle threshold^#^ | ^5^ |
| Germany | $3$ | cycle threshold^#^ | ^6^ |
| **Blood** ( $n=12$ ) |  |  |  |
| Spain | $7$ | viral load (copies/ml) | ^1^ |
| Germany | $2$ | viral load (copies/ml) | ^2^ |
| UK | $1$ | cycle threshold^#^ | ^3^ |
| Germany | $2$ | cycle threshold^#^ | ^6^ |
| **Semen** ( $n=17$ ) |  |  |  |
| Spain | $15$ | viral load (copies/ml) | ^1^ |
| Germany | $1$ | cycle threshold^#^ | ^2^ |
| Italy | $1$ | quantification cycle^#^ | ^7^ |

^#^Viral load was calculated using the conversion formula^8^.

Supplementary Table 2: Estimated fixed effect parameters, standard deviation of random effect, and standard deviation of error in mpox viral loads for each sample

| **Parameters** | **Symbol** | **Unit** | **Lesion** | | **Upper** | | **Blood** | | **Semen** | |
| --- | --- | --- | --- | --- | --- | --- | --- | --- | --- | --- |
|  |  |  | Fixed effect | SD of random effect | Fixed effect | SD of random effect | Fixed effect | SD of random effect | Fixed effect | SD of random effect |
| Clearance rate | $\delta$ | day^-1^ | $0.34$ | $0.27$ | $0.29$ | $0.40$ | $0.26$ | $0.81$ | $0.21$ | $0.21$ |
| Viral RNA loads at symptom onset | $V(0)$ | copies/ml | ${10}^{7.7}$ | $0.06$ | ${10}^{5.7}$ | $0.05$ | ${10}^{5.3}$ | $0.14$ | ${10}^{5.5}$ | $0.04$ |
| Standard deviation of error | $\sigma$ | log(copies/ml) | $1.26$ | | $1.51$ | | $0.50$ | | $1.31$ | |
